# Socioeconomic status and healthcare resource disparities among children with epilepsy in the United States: Results from a nationally representative sample

**DOI:** 10.1101/2023.05.20.23290271

**Authors:** Nallammai Muthiah, Scott Rothenberger, Taylor J. Abel

**Author notes:** **Corresponding author:** Taylor J. Abel, MD, Department of Neurological Surgery, UPMC Children’s Hospital of Pittsburgh 4401 Penn Ave, Pittsburgh, PA 15224. Declarations of interest: none.

## Abstract

**Rationale:** Epilepsy affects 1% of the US population. Healthcare and socioeconomic disparities are relatively well-studied among adults with epilepsy, but substantially less so among children. This study examines whether children with epilepsy 1) have lower SES than or 2) utilize healthcare resources differently from their peers, and 3) if SES moderates healthcare resource utilization.

**Methods:** Data from the 2016-2019 National Survey of Children’s Heath (NSCH) were used to identify children with active “epilepsy or seizure disorder”. Children with versus without epilepsy were compared. SES and healthcare utilization variables were modeled with logistic and Poisson regressions. Significance was assessed at the alpha=0.05 level.

**Results:** This analysis included 131,326 children; 835 were diagnosed with active epilepsy. The estimated population prevalence of epilepsy was 0.6%. A higher proportion of those with epilepsy were Black, non-Hispanic. Children with epilepsy were more likely to visit an ED (aOR=5.4), have seen a healthcare professional for medical care (aOR: 2.7), have ≥1 preventative checkup (aOR: 2.3), and receive medical care from a specialist (aOR: 10.3). Children from higher-income households were less likely to have epilepsy (aOR: 0.7). SES moderated the relationship between epilepsy status and ED visits. Still, 7.7% of children with epilepsy needed healthcare but did not receive it, the most common barriers being: ineligibility for services (aOR: 3.2), problems getting an appointment (aOR: 3.9), and transportation issues (aOR: 4.7).

**Conclusions:** Children with epilepsy were more likely than their peers to live in lower income households, visit EDs, and see healthcare professionals. Despite increased needs, children with epilepsy had 2.6 times the odds of not receiving needed healthcare. SES moderated the relationship between epilepsy status and healthcare resource utilization. Most common barriers to healthcare were: service eligibility, appointment scheduling, and transport. Barrier-specific policy interventions may improve medical care access for children with epilepsy.

## Introduction

Epilepsy is a common neurological condition characterized by either: 1) two unprovoked seizures occurring at least 24 hours apart, 2) one unprovoked seizure with a predisposition for further seizures, or 3) diagnosis of an epilepsy syndrome.^1^ Epilepsy affects approximately 470,000 children in the United States alone.^2^ It is estimated that one-third of these children will have seizures resistant to antiseizure medications (ASMs).^3^

Epilepsy is a chronic medical condition, and as such, health-related outcomes for people with epilepsy are influenced socioeconomic factors (Figure 1).^4^ The prevalence of epilepsy is higher among adults with low socioeconomic status (SES).^5^ At the same time, adults with lower SES are less likely to adhere to their ASM regimen.^6^ Furthermore, Black and Hispanic individuals are 30% and 40% less likely, respectively, to be seen by outpatient neurologists even after accounting for demographic, health status, and insurance differences.^7^ These statistics are alarming considering that the risk of death among people with epilepsy is up to 3-fold higher than that of the general population.^8^ Moreover, mortality risk is higher for those with poorly controlled epilepsy.^8, 9^

**Figure 1.**
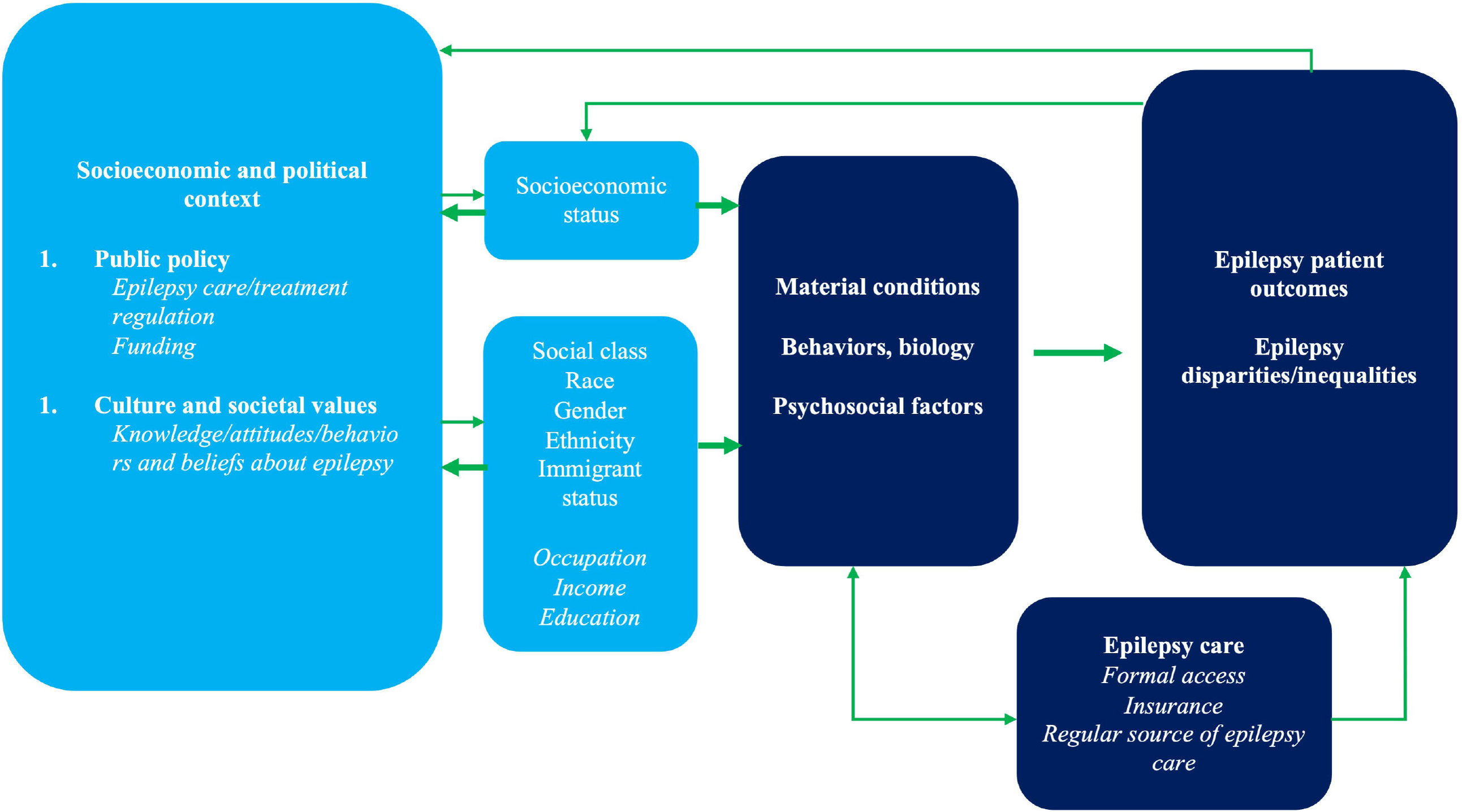
A conceptual model of the interaction between socioeconomic factors and epilepsy. Adapted from Szaflarski (2014).

While healthcare disparities are evidenced among adults with epilepsy, the extent and effect of these disparities among children remains unclear. Therefore, the primary objectives of this study were to examine whether children with epilepsy 1) have lower SES than those without epilepsy or 2) utilize healthcare resources more than those without epilepsy, and 3) if their healthcare utilization is moderated by SES. To address these objectives, we utilized four years of data from the National Survey of Children’s Heath (NSCH), a large-scale, nationally representative database addressing children’s health, family socioeconomic status, and access to medical care.

## Methods

### Sample

Data for this analysis were derived from the Maternal and Child Heath Bureau of the US Health Resources and Services Administration’s National Survey of Children’s Health (NSCH). The NSCH is a household survey which produces national and state-level data on the physical and emotional health of children 0 - 17 years old in the United States. A screening questionnaire was first administered to identify households with children as well as the number of children in the household. One child was randomly selected from each eligible household, and that child was the subject of a more detailed topical questionnaire. Responses to the topical questionnaire were weighted to represent the year’s national population. Since 2016, the NSCH has been an annual survey. To date, survey data from 2016-2019 are publicly available. Data from the 2016, 2017, 2018, and 2019 datasets were combined for the purposes of this cross-sectional analysis in accordance with the US Census Bureau’s *Guide to Multi-Year Analysis*.^10^

After combining datasets, the sample was split into groups based on the selected child’s epilepsy status: children with current, active “epilepsy or seizure disorder” versus children without active “epilepsy or seizure disorder”. Caregivers answered the question: “Has a doctor or other health care provider EVER told you that this child has epilepsy or seizure disorder?”. If the caregiver answered yes, the follow-up question was asked: “Does this child CURRENTLY have the condition?”. Therefore, response options to this question were: that the caregiver “has never been told their child has epilepsy or seizure disorder”, the caregiver “had ever been told the child has epilepsy or seizure disorder, but does not currently have the condition”, and the child “currently has epilepsy or seizure disorder”. Children who had ever been told, but do not currently have, epilepsy remained an ambiguous group. Given that seizures often occur independently from epilepsy, children who “were ever told they had epilepsy or seizure disorder” but did not identify as having active, current epilepsy were coded into the “no epilepsy” group.

### Outcome measures

Caregivers answered questions about their household income and their selected child’s healthcare resource utilization. Household income was initially collected as a continuous variable. Subsequently, household income was divided into categories for the publicly available NSCH dataset as percentage income relative to federal poverty level (FPL). Categories were: “0-99% FPL”, “100-199% FPL”, “200-399% FPL”, and “≥400% FPL”. These categories were used to represent household income.

The selected child’s number of emergency department (ED) visits over the past 12 months was used as a proxy for healthcare resource utilization. Caregivers answered the following question: “During the past 12 months, how many times did this child visit a hospital emergency room?”. Response options were: “None”, “1 time”, and “2 or more times”.

### Statistical analysis

Children with and without epilepsy were compared on caregiver and child sociodemographic measures using second-order Rao-Scott adjusted chi-square tests. Child age was compared using the adjusted Wald test. This analysis used various regression models to model outcomes of interest as a function of epilepsy status, which are described in detail below.

Ordered logistic regressions were used to determine odds ratios of 1) socioeconomic status and 2) healthcare resource utilization as a function of the selected child’s epilepsy status. As mentioned, SES was proxied by FPL categories and healthcare utilization was proxied by number of ED visits.

Multinomial logistic regressions were used to determine relative risk ratios for utilization of specific healthcare services as a function of epilepsy status. Multinomial logistic regression was used when survey item of interest contained >2 outcome categories. Multinomial logistic regressions were used to model survey items relating to: the place a child typically visits when sick and whether a child received care from a specialist healthcare provider over the last 12 months. When healthcare service variable of interest contained only two categories, a binary logistic regression was used instead to estimate odds ratios as a function of epilepsy status. Binary logistic regressions were used to model survey items relating to: whether the child saw a healthcare professional over the last 12 months, whether a child saw a healthcare professional for a preventative visit over the last 12 months, whether the child received the medical care he/she needed. In the case that needed medical care was not received, binary logistic regressions were also used to estimate odds ratios for the reason contributing to lack of needed medical care was for eligibility issues, unavailability of services, problems getting an appointment, transportation issues, the clinic/doctor’s office not being open, or cost issues as a function of epilepsy status.

Finally, a censored Poisson regression was used to estimate the incidence rate ratios of ED visits as a function of epilepsy status, adjusting for income. As mentioned, healthcare resource utilization was also modeled using ordered logistic regressions. However, given that healthcare utilization data as it existed in the dataset was right-censored (ED visits were coded in the dataset as “none”, “1”, or “2+”), censored Poisson regressions were felt to provide more accurate estimations of healthcare resource utilization. A censored Poisson regression model including an interaction between epilepsy status and income was also developed to determine if there was a moderation effect of income on the relationship between epilepsy status and healthcare resource utilization. The interaction term was tested for significance using the adjusted Wald test.

Regressions were adjusted for potential confounders. Specifically, regressions were adjusted for baseline differences between children with and without epilepsy in the baseline demographic analysis as well as differences between these children evidenced in the literature. Ultimately, all regressions were adjusted for child age and child race. For all analyses, p<0.05 was considered significant. All p-values were two-sided. All statistical analyses were performed using Stata software for Mac, Version 17.0 (StataCorp. 2021. *Stata Statistical Software: Release 17*. College Station, TX: StataCorp LLC.).

## Results

This analysis included data representing 131,326 children, among whom 835 were diagnosed with active epilepsy. The estimated population prevalence of epilepsy was 0.59% given a population size of 73,084,673. Table 1 details the demographic differences between children with versus without epilepsy. Children with epilepsy were significant older than their peers (9.2 versus 8.6 years), and a larger proportion were black, non-Hispanic (18.2% versus 13.2%). A similar proportion of children with and without epilepsy had healthcare insurance. Caregivers of children in both groups had similar levels of educational achievement.

**Table 1.**
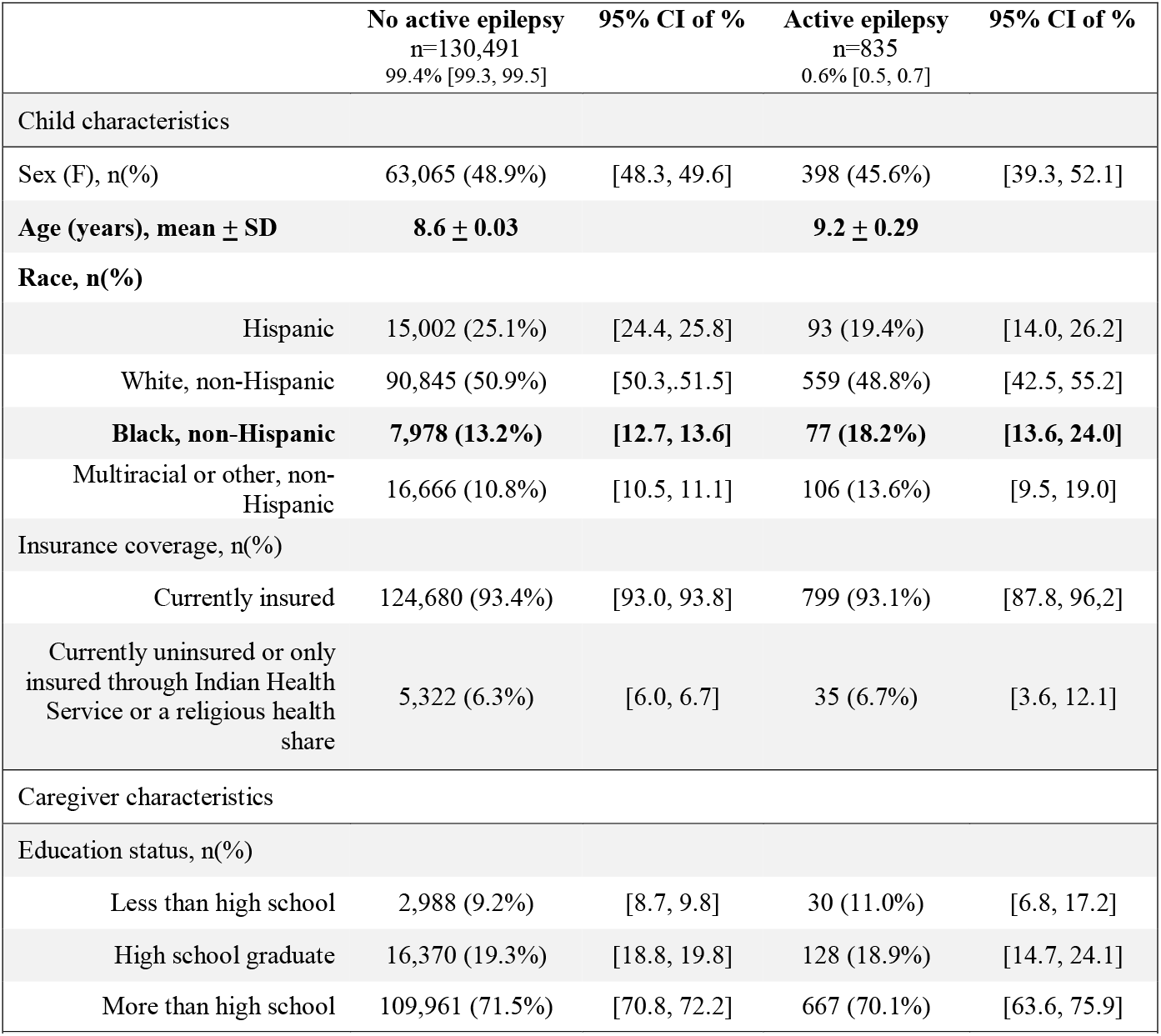
Child and caregiver characteristics of 0–17-year-old children with versus without active epilepsy from the 2016-2019 NSCH surveys (n=131,326). Prevalence figures are weighted to be nationally representative. All p-values for categorical variables reported are Pearson chi-squared values with the Rao-Scott second-order correction to account for weighted survey data; continuous outcomes were compared using the adjusted Wald test. p<0.05 was considered significant.

|  | No active epilepsy<br>n=130,491<br>99.4% [99.3, 99.5] | 95% CI of % | Active epilepsy<br>n=835<br>0.6% [0.5, 0.7] | 95% CI of % |
| --- | --- | --- | --- | --- |
| Child characteristics |  |  |  |  |
| Sex (F), n(%) | 63,065 (48.9%) | [48.3, 49.6] | 398 (45.6%) | [39.3, 52.1] |
| Age (years), mean ± SD | 8.6 ± 0.03 | | 9.2 ± 0.29 | |
| Race, n(%) |  |  |  |  |
| Hispanic | 15,002 (25.1%) | [24.4, 25.8] | 93 (19.4%) | [14.0, 26.2] |
| White, non-Hispanic | 90,845 (50.9%) | [50.3, 51.5] | 559 (48.8%) | [42.5, 55.2] |
| **Black, non-Hispanic** | **7,978 (13.2%)** | **[12.7, 13.6]** | **77 (18.2%)** | **[13.6, 24.0]** |
| Multiracial or other, non-Hispanic | 16,666 (10.8%) | [10.5, 11.1] | 106 (13.6%) | [9.5, 19.0] |
| Insurance coverage, n(%) |  |  |  |  |
| Currently insured | 124,680 (93.4%) | [93.0, 93.8] | 799 (93.1%) | [87.8, 96.2] |
| Currently uninsured or only insured through Indian Health Service or a religious health share | 5,322 (6.3%) | [6.0, 6.7] | 35 (6.7%) | [3.6, 12.1] |
| Caregiver characteristics |  |  |  |  |
| Education status, n(%) |  |  |  |  |
| Less than high school | 2,988 (9.2%) | [8.7, 9.8] | 30 (11.0%) | [6.8, 17.2] |
| High school graduate | 16,370 (19.3%) | [18.8, 19.8] | 128 (18.9%) | [14.7, 24.1] |
| More than high school | 109,961 (71.5%) | [70.8, 72.2] | 667 (70.1%) | [63.6, 75.9] |

Table 2 depicts the odds ratios associated with healthcare resource utilization (proxied as number of ED visits) and socioeconomic status (proxied as percentage of income relative to FPL) for those with versus without epilepsy. Ordered logistic regression demonstrated that children who visited the ED more times had incrementally higher odds of having active epilepsy. Children with epilepsy (compared to those without) had 5.4 times the adjusted odds of 2+ ED visits than 0-1 ER visits. Similarly, the adjusted odds of 1-2+ ER visits was 5.4 times the odds of 0 ER visits for children with epilepsy compared to those without. Children in lower-income groups also had incrementally higher odds of having active epilepsy. Regression analysis demonstrated that children with epilepsy (compared to those without) had 0.65 times the adjusted odds of an income ≥400% FPL than anything less. Similarly, the adjusted odds of having an income of the higher income categories (200-399% FPL or ≥400%% FPL) were 0.65 times that of the lower income categories (0-99% FPL or 100-199% FPL) for those with epilepsy. By extension, the odds of an income 100-199% FPL or higher were 0.65 times the odds of an income 0-99% for children with epilepsy.

**Table 2.** Univariable logistic regression models detailing the number of emergency department visits and average household income (as % of federal poverty level) among children with versus without active epilepsy from the 2016-19 NSCH (n=131,326). OR: odds ratio; CI: confidence interval; aOR: adjusted odds ratio. ^*^aOR adjusted for child age and child race. Results are weighted to be nationally representative. Significance was assessed at the 0.05 level.

|  | No active epilepsy<br>n=130,491<br>99.4% [99.3, 99.5] | 95% CI of<br>% | Active epilepsy<br>n=835<br>0.6% [0.5, 0.7] | 95% CI of<br>% | OR<br>[95% CI] | aOR*<br>[95% CI] |
| --- | --- | --- | --- | --- | --- | --- |
| Number of emergency department visits, n(%) |  |  |  |  |  |  |
| None | 107,871 (80.5%) | [79.9, 81.0] | 449 (48.7%) | [42.3, 55.2] | ref | ref |
| 1 | 17,566 (15.0%) | [14.6, 15.5] | 198 (25.2%) | [20.2, 31.1] | **2.77 [2.03-3.79]** | **2.91 [2.13-3.98]** |
| 2+ | 4,535 (4.5%) | [4.3, 4.8] | 185 (26.1%) | [20.7, 32.2] | **9.53 [6.87-13.21]** | **10.17 [7.28-14.20]** |
| Household income, % of federal poverty level, n(%) |  |  |  |  |  |  |
| 0-99% | 14,347 (20.2%) | [19.6, 20.8] | 142 (30.1%) | [23.9, 37.2] | ref | ref |
| 100-199% | 20,878 (21.7%) | [21.1, 22.2] | 171 (20.9%) | [16.7, 25.9] | **0.65 [0.45-0.94]** | **0.65 [0.45-0.94]** |
| 200-399% | 40,357 (27.5%) | [27.0, 28.0] | 229 (23.5%) | [18.5, 29.2] | **0.57 [0.39-0.85]** | **0.56 [0.37-0.83]** |
| ≥400% | 54,909 (30.6%) | [30.1, 31.1] | 293 (25.5%) | [20.8, 30.9] | **0.56 [0.39-0.80]** | **0.53 [0.36-0.77]** |

Table 3 demonstrates the odds ratios or relative risk ratios for utilization of various medical services/resources between children without and without epilepsy. Overall, compared to their peers, children with epilepsy were more likely to attend preventative medicine visits and less likely to receive needed healthcare. Specifically, 93% of children with epilepsy and 83% of children without epilepsy had seen a healthcare professional for medical care over the last year. Children with epilepsy had 2.7 times the adjusted odds of seeing any healthcare professional for medical care over the last 12 months and 2.3 times the adjusted odds of seeing a healthcare professional for at least one preventative checkup over the last 12 months than their peers. When sick, children with epilepsy had 4.7 times the relative risk of going to a hospital ED (compared to a doctor’s office) and 2.5 times the relative risk of going to a hospital outpatient department (compared to a doctor’s office) than children without epilepsy. Overall, 61% of children with epilepsy received care from a specialist doctor other than a mental health profession compared to only 14% of their peers (aOR: 10.3, CI: 7.7-13.8). Children with epilepsy had 2.6 times the adjusted odds of not receiving needed healthcare than children without (8% versus 3%, respectively). Reasons contributing to why children did not receive needed healthcare services included that the child was not eligible for the needed healthcare services (aOR: 3.2, CI: 1.0[2]-10.2), problems getting an appointment (aOR: 3.9, CI: 2.4-6.4), and transportation issues (aOR: 4.7, CI: 2.0-11.0).

**Table 3.** Utilization of medical services among children with versus without active epilepsy from the 2016-19 NSCH (n=131,326). OR: odds ratio; RRR: relative risk ratio; CI: confidence interval; aOR: adjusted odds ratio; aRRR: adjusted relative risk ratio; NE: not estimatable. *aOR and aRRR adjusted for child age and child race. Results are weighted to be nationally representative. Significance was assessed for adjusted analyses at the 0.05 level. ^+^reference: ȜDid not experience this difficulty in receiving needed medical careȝ

|  | No active epilepsy<br>n=130,491<br>99.4%<br>[99.3, 99.5] | 95% CI of % | Active epilepsy<br>n=835<br>0.6%<br>[0.5, 0.7] | 95% CI of % | OR or RRR<br>(95% CI) | aOR* or aRRR*<br>(95% CI) |
| --- | --- | --- | --- | --- | --- | --- |
| Child saw a healthcare professional for medical care over the last 12 months |  |  |  |  |  |  |
| No | 17,443 (17.0%) | [16.5, 17.6] | 30 (7.2%) | [4.0, 12.8] | ref | ref |
| Yes | 112,894 (83.0%) | [82.5, 83.5] | 805 (92.8%) | [87.2, 96.0] | **2.63 [1.39, 4.97]** | **2.69 [1.42, 5.10]** |
| Child saw a healthcare professional for at least one preventative checkup over the last 12 months |  |  |  |  |  |  |
| No | 22,111 (20.4%) | [19.9, 21.0] | 62 (10.1%) | [6.4, 15.5] | ref | ref |
| Yes | 107,504 (79.6%) | [79.0, 80.1] | 767 (90.0%) | [84.5, 93.6] | **2.29 [1.40, 3.77]** | **2.34 [1.43, 3.87]** |
| Place the child usually goes when sick |  |  |  |  |  |  |
| Doctor's office | 94,422 (75.0%) | [74.4, 75.7] | 620 (74.4%) | [67.2, 80.5] | ref | ref |
| Hospital emergency room | 799 (1.4%) | [1.2, 1.6] | 29 (6.8%) | [3.9, 11.7] | **4.92 [2.67, 9.06]** | **4.73 [2.44, 9.18]** |
| Hospital outpatient department | 549 (0.6%) | [0.5, 0.8] | 15 (1.7%) | [0.7, 3.8] | **2.62 [1.09, 6.29]** | **2.52 [1.04, 6.11]** |
| Clinic/health center | 10,109 (8.3%) | [7.9, 8.7] | 49 (7.0%) | [3.4, 14.1] | 0.85 [0.39, 1.86] | 0.85 [0.40, 1.81] |
| Minute clinic | 950 (0.8%) | [0.7, 0.8] | 3 (0.5%) | [0.1, 1.7] | 0.64 [0.18, 2.26] | 0.61 [0.17, 2.13] |
| School | 601 (0.4%) | [0.3, 0.4] | 6 (0.6%) | [0.2, 1.9] | 1.59 [0.49, 5.22] | 1.47 [0.45, 4.79] |
| Somewhere else | 588 (0.5%) | [0.4, 0.6] | 4 (0.8%) | [0.2, 3.5] | 1.61 [0.34, 7.67] | 1.57 [0.33, 7.44] |
| Received care from a specialist doctor other than a mental health professional during the past 12 months |  |  |  |  |  |  |
| No, and he/she needed to | 106,109 (84.5%) | [84.1, 84.9] | 230 (37.3%) | [31.1, 43.9] | ref | ref |
| No, but he/she needed to | 1,869 (1.8%) | [1.6, 2.0] | 11 (1.8%) | [0.9, 3.8] | **2.29 [1.04, 5.03]** | 2.15 [0.98, 4.75] |
| Yes | 21,502 (13.7%) | [13.3, 14.1] | 586 (60.9%) | [54.3, 67.1] | **10.05 [7.61, 13.27]** | **10.32 [7.73, 13.77]** |
| Any times when child needed healthcare but it was not received |  |  |  |  |  |  |
| Received needed care | 126,823 (96.6%) | [96.3, 96.9] | 775 (92.2%) | [87.5, 95.2] |  |  |
| Needed healthcare not received | 3,200 (3.0%) | [2.8, 3.3] | 59 (7.7%) | [4.6, 12.4] | **2.63 [1.54, 4.52]** | **2.58 [1.49, 4.46]** |
| Reasons contributing to why child did not receive needed health services: <sup>+</sup> |  |  |  |  |  |  |
| Child was not eligible | 863 (1.0%) | [0.9, 1.1] | 16 (3.0%) | [1.0, 8.8] | **3.25 [1.04, 10.08]** | **3.22 [1.02, 10.15]** |
| Services were not available in area | 743 (0.7%) | [0.6, 0.8] | 22 (1.2%) | [1.0, 3.3] | NE | NE |
| Problems getting an appointment | 1,213 (1.2%) | [1.0, 1.3] | 39 (4.5%) | [2.9, 7.0] | **4.13 [2.54, 6.71]** | **3.94 [2.41, 6.44]** |
| Transportation issues | 359 (0.4%) | [0.3, 0.5] | 9 (1.9%) | [0.8, 4.3] | **4.77 [2.02, 11.23]** | **4.67 [1.99, 10.96]** |
| Clinic/doctor's office was not open | 362 (0.4%) | [0.3, 0.5] | 8 (1.0%) | [0.4, 2.7] | **2.80 [1.01, 7.74]** | 2.72 [0.99, 7.46] |
| Cost issues | 1,956 (1.8%) | [1.6, 2.0] | 25 (2.3%) | [1.3, 4.2] | 1.34 [0.72, 2.51] | 1.34 [0.72, 2.51] |

Table 4 demonstrates the results of the censored Poisson regression to assess healthcare resource utilization in this sample. The incidence rate of visiting the ED children was 2.6 times higher for children with epilepsy than for children without. As children’s income category increased, their adjusted incidence rate of visiting the ED decreased (IRR for 100-199% FPL: 0.73, CI: 0.67-0.80; IRR for 200-399% FPL: 0.55, CI: 0.50-0.60; IRR for ≥400% FPL: 0.48, CI: 0.45-0.52). The overall interaction term between epilepsy status and household income category was found to be significant (p=0.02). Post-hoc analysis of the interaction revealed that the incidence rate ratio associated with epilepsy is 2.04 times higher for children in the 200-399% FPL income category compared to those in the 0-99% FPL income category (IRR 2.04, CI: 1.28-3.24). In other words, the effect of epilepsy on ED visit incidence rate is twice as large for children in the second-to-highest income category compared to those in the lowest income category.

**Table 4.** Censored Poisson regression to assess healthcare utilization (number of emergency room visits over the last 12 months) among children with versus without active epilepsy. Data are from the 2016-19 NSCH (n=131,326). IRR: incident rate ratio; CI: confidence interval; aIRR: adjusted incident rate ratio. ^*^aIRR adjusted for child age and child race. Results are weighted to be nationally representative. Significance was assessed for the adjusted analysis at the 0.05 level.

|  | **IRR**<br>**[95% CI]** | **aIRR\***<br>**[95% CI]** |
| --- | --- | --- |
| Active epilepsy | **2.52 [1.80-3.52]** | **2.56 [1.84-3.57]** |
| Household income, % of federal poverty level, n(%) |  |  |
| 0-99% | ref | ref |
| 100-199% | **0.70 [0.64-0.76]** | **0.73 [0.67-0.80]** |
| 200-399% | **0.51 [0.47-0.55]** | **0.55 [0.50-0.60]** |
| ≥400% | **0.44 [0.40-0.47]** | **0.48 [0.45-0.52]** |
| Interaction between Household income, % of federal poverty level, n(%) and epilepsy status |  |  |
| 0-99% x no epilepsy | ref | ref |
| 100-199% x epilepsy | 1.28 [0.79-2.07] | 1.34 [0.84-2.16] |
| 200-399% x epilepsy | **2.04 [1.28-3.27]** | **2.04 [1.28-3.24]** |
| ≥400% x epilepsy | 1.30 [0.84-2.03] | 1.29 [0.83-2.02] |

## Discussion

We performed a cross-sectional analysis of the socioeconomic factors and healthcare resource utilization among children with versus without epilepsy in the United States, using four years of nationally representative data. In a sample of 131,326 children, our results demonstrate that children with epilepsy had significantly higher odds of: having lower household income, visiting the ED more frequently, and not receiving needed healthcare. We also found that SES (proxied by percentage of FPL) moderated the effect of epilepsy status on healthcare resource utilization (proxied by ED visits).

From this weighted sample, we estimated the national prevalence of epilepsy to be 0.59%. Scaled to the US population (estimated size: 73,084,673), this represents approximately 431,200 children, which is slightly lower than the current estimate of 470,000.^2^ We expect that some children in the sample, especially those who “were ever told they had epilepsy or seizure disorder”, may actually have had epilepsy, leading to potential underrepresentation of the true population of US children with epilepsy.

The first objective of this analysis was to assess whether US children with epilepsy had lower SES than those without. Such a relationship has been well-evidenced in US adult and non-US populations. A recent meta-analysis by Fiest et al. demonstrated that the active annual period prevalence of epilepsy was higher in low-to-middle income countries than high-income countries.^11^ Regarding differences in SES among patients within individual countries, Noronha et al. demonstrated in a Brazilian population that (even after adjusting for differences in treatment) there was a higher prevalence of epilepsy among lower-income SES patients.^12^ In Sweden, Li et al. revealed that lower education status and lower income were both associated with increased risk of hospitalization for epilepsy.^13^ Evidence from a national study in Iceland suggests that the risk of epilepsy is higher in adults with low socioeconomic status, but that the same relationship does not exist for children.^5^ Moreover, in Zambia, Birbeck et al. found that people with epilepsy not only had lower education status, but also poorer living conditions than those without epilepsy.^14^ SES has been evidenced to profoundly influence treatment adherence, morbidity, and mortality among epilepsy patients.^5, 6, 8, 9^

The above-mentioned findings reflect non-US or primarily adult populations. Evidence for an association between epilepsy and SES in children is mixed^15^, yet lower SES in epilepsy has been associated with delayed care and potentially worse outcomes in children with epilepsy from other developed countries.^16, 17^ Our results in an exclusively US pediatric sample demonstrated that even after adjusting for age and race, children with epilepsy had 1.47 times the odds (the reciprocal of OR=0.68, as presented) of having lower income than their peers. The socioeconomic status of children in this analysis is likely more a reflection of the socioeconomic status of their caregivers. Given that an estimated 70-80% of epilepsy is genetic^18, 19^, it is possible that some children in this analysis with epilepsy have a caregiver with epilepsy. The average total direct healthcare cost of epilepsy per person in the United States is estimated to range from $10,000-$48,000.^20^ Coupled with evidence that adults with epilepsy are more likely to be unemployed or unable to work and have comorbid health conditions^21^ which may pose additional barriers in accessing necessary treatment for their children, a “downward socioeconomic spiral” may exist. Thus, it is possible that children with epilepsy in our study may have had lower SES because their caregivers also had epilepsy themselves. Further work would need to investigate this hypothesis.

Our second objective was to assess whether children with epilepsy utilized medical resources more than those without epilepsy. Existing literature suggests that adults living with epilepsy and lower SES have higher healthcare utilization and concomitantly worse epilepsy outcomes.^22^ Children with epilepsy had 5.4 times the incidence rate of 2+ ED visits than 0-1 visits than their peers. The NSCH data does not offer information on the reason for these increased ED visits. However, it is likely that many of these additional ED visits were related to epilepsy: increased seizure activity, injuries related to seizure activity, or for emergency antiseizure medication-related adverse events. Seizures and epilepsy care are the most common neurological reason for presentation to an ED.^23^ Visiting the ED is economically- and psychologically-taxing for patients and caregivers. It has further been argued that many ED visits for epilepsy could be avoided.^24^ Ryan et al. found that the financial cost associated with pediatric epilepsy is highest (around $20,000) the first year after diagnosis, and that ED visits comprise the third-highest cost (second only to hospital admissions and diagnostic procedures) while only accounting for 1% of total visits.^24^ Patel et al. demonstrated that a targeted quality improvement intervention (focused on five key interventions: establishing an urgent epilepsy clinic, improving at-home seizure management plans, making information on proper abortive seizure medication dosing more accessible, reminder magnets with information on abortive seizure medications, and targeting unique issues for patients who tended to use the ED frequently) reduced ED visits by 28% over 19 months.^25^

Crucially, our analysis suggests that children with epilepsy are 2.6 more likely to not receive needed healthcare. The primary reasons cited for lack of needed care were: the child was not eligible for needed healthcare services, caregivers had difficulty getting an appointment, and transportation was a prohibitive factor. Lack of adequate treatment for epilepsy has been associated with worse epilepsy outcomes including higher mortality rates in those with untreated epilepsy.^26^ These factors are similar to those targeted by Patel et al. in their single-center quality improvement project. The results of analysis provide meaningful potential targets health policy targets to improve access to epilepsy care for US children. Our study demonstrated that transportation barriers are the most burdensome for caregivers. That said, the results of this analysis reflect the averaged national population. Each community is unique in its barriers to healthcare. Solomon et al. in a recent systematic review, suggest that making transportation more affordable or easy-to-access, while important, may not be sufficient by itself to improve access to needed healthcare resources.^27^ Thus, community-based, institutional- or state-level targeted quality improvement interventions, like that performed by Patel et al., may be the most effective way to identify and target the unique factors contributing to lack of access to needed healthcare.

The final objective of this analysis was to determine whether the association of epilepsy with healthcare resource utilization was moderated by SES. Unlike some other developed countries^17^, the US lacks universal healthcare or centralized epilepsy care delivery mechanisms. Our results revealed evidence that the effect of having active epilepsy on ED usage may differ by income category. Evidence in adults samples indicates that some of the primary drivers of the moderation effect of SES on healthcare resource utilization are differences in health at baseline.^28-31^ Our analysis focused on children, who likely have similar baseline health status to each other compared with adults. Perhaps children with epilepsy in higher income groups have fewer limitations in access to EDs than children at or below FLP. In conjunction, they may have more events inciting EDs visits than children without epilepsy. Still, the interaction between income category and epilepsy status was not incremental, as expected. Thus, the results of this moderation analysis provide evidence of a potential interaction between epilepsy status and SES on healthcare resource utilization, but the true nature and underlying reasons for this effect require further investigation.

### Limitations and Future Work

This study has several limitations. First, it is cross-sectional in nature, and is limited by biases which limit all cross-sectional studies. No inferences about causation, only correlations and associations, can be drawn from our results. That said, the data used for this analysis reflected over 131,000 children in the US and were weighted to reflect the US population. This analysis also extended only to 2019 due to availability of data. Considering the significant socioeconomic changes brought forth by the global pandemic, it will become critical to re-characterize the state of SES and healthcare utilization among children with epilepsy in the post-pandemic era so that specific local-, state-, and federal-level interventions can be designed to improve access to epilepsy care. Finally, this analysis used simple proxies for complex socioeconomic concepts. These proxies were chosen based on precedents set in prior literature, based on data availability, and due to lack of standardized and validated methodology for utilizing base survey data to derive complex composite variables like SES or healthcare resource utilization. If future studies were able to validate such derivation methods for use in large-scale dataset analysis, researchers would be able to make even better use of the rich data available from well-performed cross-sectional studies like the NSCH, and more comprehensive, nuanced characterizations of complex socioeconomic constructs would be possible.

## Conclusions

Children with epilepsy are more likely than their peers to live in lower income households, visit an ED, and see healthcare professionals. Income category may moderate the relationship between epilepsy status and healthcare resource utilization. Children with active epilepsy had 2.6 times the odds of not receiving necessary healthcare, with the most common barriers being: service eligibility, appointment scheduling, and transport. Health policy interventions to alleviate these barriers will improve access to needed medical care for children with epilepsy specifically, and for those with other chronic medical conditions broadly.

## Data Availability

Source data are openly available to the public.

## References

1. Fisher RS, Acevedo C, Arzimanoglou A, et al. ILAE official report: a practical clinical definition of epilepsy. Epilepsia. Apr 2014;55(4):475–82. doi:10.1111/epi.12550

2. Zack MM, Kobau R. National and State Estimates of the Numbers of Adults and Children with Active Epilepsy -United States, 2015. MMWR Morb Mortal Wkly Rep. Aug 11 2017;66(31):821–825. doi:10.15585/mmwr.mm6631a1

3. Kalilani L, Sun X, Pelgrims B, Noack-Rink M, Villanueva V. The epidemiology of drugresistant epilepsy: A systematic review and meta-analysis. Epilepsia. Dec 2018;59(12):2179–2193. doi:10.1111/epi.14596

4. Szaflarski M. Social determinants of health in epilepsy. Epilepsy Behav. Dec 2014;41:283–9. doi:10.1016/j.yebeh.2014.06.013

5. Hesdorffer DC, Tian H, Anand K, et al. Socioeconomic status is a risk factor for epilepsy in Icelandic adults but not in children. Epilepsia. Aug 2005;46(8):1297–303. doi:10.1111/j.1528-1167.2005.10705.x

6. Burneo JG, Jette N, Theodore W, et al. Disparities in epilepsy: report of a systematic review by the North American Commission of the International League Against Epilepsy. Epilepsia. Oct 2009;50(10):2285–95. doi:10.1111/j.1528-1167.2009.02282.x

7. Saadi A, Himmelstein DU, Woolhandler S, Mejia NI. Racial disparities in neurologic health care access and utilization in the United States. Neurology. Jun 13 2017;88(24):2268–2275. doi:10.1212/WNL.0000000000004025

8. Ficker DM. Sudden unexplained death and injury in epilepsy. Epilepsia. 2000;41 Suppl 2:S7–12. doi:10.1111/j.1528-1157.2000.tb01519.x

9. Nilsson L, Farahmand BY, Persson PG, Thiblin I, Tomson T. Risk factors for sudden unexpected death in epilepsy: a case-control study. Lancet. Mar 13 1999;353(9156):888–93. doi:10.1016/s0140-6736(98)05114-9

10. Bureau UC. Guide to Multi-Year Analysis. Commerce UDo; 2021. https://www2.census.gov/programs-surveys/nsch/technical-documentation/methodology/NSCH-Guide-to-Multi-Year-Estimates.pdf

11. Fiest KM, Sauro KM, Wiebe S, et al. Prevalence and incidence of epilepsy: A systematic review and meta-analysis of international studies. Neurology. Jan 17 2017;88(3):296–303. doi:10.1212/WNL.0000000000003509

12. Noronha AL, Borges MA, Marques LH, et al. Prevalence and pattern of epilepsy treatment in different socioeconomic classes in Brazil. Epilepsia. May 2007;48(5):880–5. doi:10.1111/j.1528-1167.2006.00974.x

13. Li X, Sundquist J, Sundquist K. Socioeconomic and occupational risk factors for epilepsy: a nationwide epidemiological study in Sweden. Seizure. Apr 2008;17(3):254–60. doi:10.1016/j.seizure.2007.07.011

14. Birbeck G, Chomba E, Atadzhanov M, Mbewe E, Haworth A. The social and economic impact of epilepsy in Zambia: a cross-sectional study. Lancet Neurol. Jan 2007;6(1):39–44. doi:10.1016/S1474-4422(06)70629-9

15. Durkin MS, Yeargin-Allsopp M. Socioeconomic Status and Pediatric Neurologic Disorders: Current Evidence. Semin Pediatr Neurol. Oct 2018;27:16–25. doi:10.1016/j.spen.2018.03.003

16. Geerts A, Brouwer O, van Donselaar C, et al. Health perception and socioeconomic status following childhood-onset epilepsy: the Dutch study of epilepsy in childhood. Epilepsia. Dec 2011;52(12):2192–202. doi:10.1111/j.1528-1167.2011.03294.x

17. Puka K, Smith ML, Moineddin R, Snead OC, Widjaja E. The influence of socioeconomic status on health resource utilization in pediatric epilepsy in a universal health insurance system. Epilepsia. Mar 2016;57(3):455–63. doi:10.1111/epi.13290

18. Kjeldsen MJ, Corey LA, Christensen K, Friis ML. Epileptic seizures and syndromes in twins: the importance of genetic factors. Epilepsy Res. Jun-Jul 2003;55(1-2):137–46. doi:10.1016/s0920-1211(03)00117-7

19. Myers CT, Mefford HC. Advancing epilepsy genetics in the genomic era. Genome Med. Aug 25 2015;7:91. doi:10.1186/s13073-015-0214-7

20. Begley CE, Durgin TL. The direct cost of epilepsy in the United States: A systematic review of estimates. Epilepsia. Sep 2015;56(9):1376–87. doi:10.1111/epi.13084

21. Kobau R, Zahran H, Thurman DJ, et al. Epilepsy surveillance among adults--19 States, Behavioral Risk Factor Surveillance System, 2005. MMWR Surveill Summ. Aug 8 2008;57(6):1–20.

22. Groover O, Morton ML, Janocko NJ, et al. Mind the gap: health disparities in families living with epilepsy are significant and linked to socioeconomic status. Epileptic Disord. Dec 1 2020;22(6):782–789. doi:10.1684/epd.2020.1229

23. Pallin DJ, Goldstein JN, Moussally JS, Pelletier AJ, Green AR, Camargo CA, Jr. Seizure visits in US emergency departments: epidemiology and potential disparities in care. Int J Emerg Med. Jun 2008;1(2):97–105. doi:10.1007/s12245-008-0024-4

24. Ryan JL, McGrady ME, Guilfoyle SM, Junger K, Arnett AD, Modi AC. Health care charges for youth with newly diagnosed epilepsy. Neurology. Aug 11 2015;85(6):490–7. doi:10.1212/WNL.0000000000001746

25. Patel AD, Wood EG, Cohen DM. Reduced Emergency Department Utilization by Patients With Epilepsy Using QI Methodology. Pediatrics. Feb 2017;139(2)doi:10.1542/peds.2015-2358

26. Beghi E. The Epidemiology of Epilepsy. Neuroepidemiology. 2020;54(2):185–191. doi:10.1159/000503831

27. Solomon EM, Wing H, Steiner JF, Gottlieb LM. Impact of Transportation Interventions on Health Care Outcomes: A Systematic Review. Med Care. Apr 2020;58(4):384–391. doi:10.1097/MLR.0000000000001292

28. Droomers M, Westert GP. Do lower socioeconomic groups use more health services, because they suffer from more illnesses? Eur J Public Health. Sep 2004;14(3):311–3. doi:10.1093/eurpub/14.3.311

29. Loef B, Meulman I, Herber GM, et al. Socioeconomic differences in healthcare expenditure and utilization in The Netherlands. BMC Health Serv Res. Jul 3 2021;21(1):643. doi:10.1186/s12913-021-06694-9

30. Meulman I, Uiters E, Polder J, Stadhouders N. Why does healthcare utilisation differ between socioeconomic groups in OECD countries with universal healthcare coverage? A protocol for a systematic review. BMJ Open. Nov 23 2021;11(11):e054806. doi:10.1136/bmjopen-2021-054806

31. van Doorslaer E, Wagstaff A, van der Burg H, et al. Equity in the delivery of health care in Europe and the US. J Health Econ. Sep 2000;19(5):553–83. doi:10.1016/s0167-6296(00)00050-3

